# Expanding the genotype and phenotype spectrum of *SYT1*-associated neurodevelopmental disorder

**DOI:** 10.1101/2021.07.21.21260857

**Authors:** Holly Melland, Fabian Bumbak, Anna Kolesnik-Taylor, Elise Ng-Cordell, Abinayah John, Panayiotis Constantinou, Shelagh Joss, Martin Larsen, Christina Fagerberg, Jenny Thies, Frances Emslie, Marjolein Willemsen, Tjitske Kleefstra, Rolf Pfundt, Rebekah Barrick, Richard Chang, Lucy Loong, Majid Alfadhel, Jasper van der Smagt, Mathilde Nizon, Manju Kurian, Daniel J Scott, Joshua J Ziarek, Sarah Gordon, Kate Baker

**Affiliations:** The Florey Institute of Neuroscience and Mental Health, University of Melbourne, Parkville, VIC, Australia; Melbourne Dementia Research Centre, The Florey Institute of Neuroscience and Mental Health, University of Melbourne, Parkville, VIC, Australia; Department of Molecular and Cellular Biochemistry, Indiana University, Bloomington, IN, USA; MRC Cognition and Brain Sciences Unit, University of Cambridge, Cambridge, UK; Department of Clinical Genetics, Queen Elizabeth University Hospital, Glasgow, UK; Odense University Hospital, Odense, Denmark; Department of Pediatrics, Division of Genetic Medicine, Seattle Children’s Hospital, Seattle, WA, USA; South West Thames Regional Genetics Service at St George’s, University of London, London, UK; Radboud University Medical Center, Nijmegen, NL; Vincent van Gogh Centre for Neuropsychiatry, Venray, NL; Children’s Hospital of Orange County, Orange, CA, USA; Oxford Centre for Genomic Medicine, Oxford University Hospitals NHS Foundation Trust, Oxford, UK; Genetics and Precision Medicine department, King Abdullah Specialized Children Hospital, King Abdulaziz Medical City, Ministry of National Guard Health Affairs, Riyadh, Saudi Arabia; Medical Genomics Research Department, King Abdullah International Medical Research Center, Ministry of National Guard Health Affairs, Riyadh, Saudi Arabia; College of Medicine, King Saud bin Abdulaziz University for Health Sciences, King Abdulaziz Medical City, Ministry of National Guard Health Affairs, Riyadh, Saudi Arabia; Utrecht University Medical Centre, Utrecht, NL; CHU Nantes, Service de Génétique Médicale, INSERM, Université de Nantes, Nantes, France; Developmental Neurosciences Programme, UCL Institute of Child Health, London, UK; Department of Medical Genetics, University of Cambridge, Cambridge, UK

**Author notes:** <u>Corresponding Author</u> Correspondence to Kate Baker.

## Abstract

**Purpose:** Synaptotagmin-1 (SYT1) is a critical mediator of neurotransmitter release in the central nervous system. Previously reported missense *SYT1* variants in the C2B domain are associated with severe intellectual disability, movement disorders, behavioural disturbance and EEG abnormalities. Here, we expand the genotypes and phenotypes and identify discriminating features of this disorder.

**Methods:** We describe 22 individuals with 15 *de novo* missense *SYT1* variants. Evidence for pathogenicity is discussed, including ACMG criteria, known structure-function relationships, and molecular dynamics simulations. Quantitative behavioural data for 14 cases are compared to other monogenic neurodevelopmental disorders.

**Results:** Four variants lie in the C2A domain with the remainder in the C2B. We classify 6 variants as pathogenic, 4 as likely pathogenic, and 5 as variants of uncertain significance. Prevalent clinical phenotypes include delayed developmental milestones, ophthalmic problems, abnormal EEG, movement disorders and sleep disturbance. Discriminating behavioural characteristics were severity of motor and communication impairment, presence of motor stereotypies and mood instability.

**Conclusion:** *SYT1* variants associated with neurodevelopmental disorder extend beyond previously reported regions, and the phenotypic spectrum encompasses a broader range of severity than initially reported. This work guides diagnosis and molecular understanding of this rare neurodevelopmental disorder, and highlights a key role for SYT1 function in emotional regulation, motor control and emergent cognitive function.

## Introduction

The tightly regulated synaptic vesicle cycle involves the trafficking, docking, fusion, and recycling of neurotransmitter-filled vesicles at the presynaptic terminal. Precision and efficiency of these processes is critical for synchronous neurotransmission, neural network development and emergent cognitive functions^1,2^. Inherited and *de novo* variants in synaptic vesicle cycling genes have been associated with a broad spectrum of neurodevelopmental phenotypes including epilepsies, movement disorders, delayed acquisition of motor milestones, intellectual disability (ID), visual impairment and emotional-behavioural disturbances^3^. Improving the delineation, diagnosis, and management of these disorders requires comprehensive phenotyping in parallel with detailed genetic, molecular and cellular analysis of variants.

Variants in the *SYT1* gene, coding for synaptotagmin-1 (SYT1) give rise to *SYT1*-associated neurodevelopmental disorder, also known as Baker-Gordon Syndrome (MIM 618218). SYT1 is a synaptic vesicle protein that couples action potentials to the synchronous exocytosis of neurotransmitters through its calcium sensing activity^4^. SYT1, the dominant synaptotagmin family member in the forebrain^5,6^, is a transmembrane protein with two cytoplasmic calcium-binding (C2A and C2B) domains^7^. Membrane depolarisation triggers an influx of Ca^2+^ ions into the nerve terminal, which bind to negatively charged aspartate residues that reside in loops at the “top” of each C2 domain. This neutralises the charge of the loops, acting as an electrostatic switch and allowing the hydrophobic tips of these loops to penetrate the negatively-charged plasma membrane, thereby facilitating fusion of synaptic vesicle and plasma membranes^4^.

We previously reported 11 cases of *de novo* missense variants in *SYT1*^8,9^. All affected individuals presented with hypotonia, developmental delay, and ID varying from moderate to profound severity, and one-third exhibited symptoms of an involuntary movement disorder. Behavioural characteristics included unpredictable switches from placidity to agitation, and pronounced motor stereotypies such as hand-biting. EEG was abnormal in all cases, characterised by intermittent low frequency high amplitude oscillations. Five different *SYT1* variants were identified (Met303Lys, Asp304Gly, Asp366Glu, Ile368Thr, and Asn371Lys), all located in highly conserved residues of the C2B domain of SYT1, clustering around the Ca^2+^-binding pocket. These missense variants were found to inhibit evoked exocytosis in a dominant-negative and mutation-specific manner^9,10^.

The present paper expands the genotypic and phenotypic spectrum of *SYT1*-associated neurodevelopmental disorder. We evaluate evidence for pathogenicity of novel variants through *in silico* analysis and molecular dynamics simulations. We provide quantitative evaluation of behavioural characteristics within the cohort and comparison to individuals with other monogenic neurodevelopmental disorders.

## Materials and Methods

### Recruitment and sample description

*SYT1* variants were identified via whole exome sequencing (trio or solo) within clinical laboratories or ethically approved research studies. The identification, validation, confirmation of *de novo* status, and clinical reporting of *SYT1* variants was carried out by each participant’s clinical centre. Notification of diagnosed variants was communicated to senior authors of the study by personal communication, or identified through database searching (ClinVar, Decipher) or GeneMatcher. An additional 51 participants with ID of known monogenic origin (excluding synaptic vesicle cycling disorders as listed by John et al. (2021)^3^) were recruited as a comparison group for behavioural data. Genetic diagnoses within the comparison group are listed in Supplementary Materials (Table S1). The SYT1 and ID comparison groups did not significantly differ in range and distributions of age, sex, and global adaptive function (Table S2).

### Evaluation of variants

Evaluation of pathogenicity for the 15 *de novo* sequence variants followed the American College of Medical Genetics and Genomics and the Association for Molecular Pathology (ACMG) classification guidelines^11^ supplemented by Association for Clinical Genomic Science (ACGS) UK best practice guidelines^12^ (for details see Supplementary Material).

### Molecular Dynamics Simulations

Homology models of mutant versions of SYT1 C2 domains were generated from Ca^2+^-bound C2A and C2B solution structures (PDB: 1BYN^13^ and 1K5W^14^; note that amino acid numbering used throughout this paper follows human sequence for simplicity). The WT structure and each mutated homology model (with Ca^2+^ ions both present and removed), were subjected to four individual atomistic molecular dynamics simulations with trajectory lengths of ∼400ns each (for full details see Supplementary Material). Root-mean-square deviation (RMSD) of the backbone and root-mean-square fluctuations (RMSF) of the backbone C-alpha atoms of each domain variant were measured over the course of the simulations as readouts of overall and local mobility of the domains. To assess Ca^2+^ mobility and binding pocket occupancy, the distance between each Ca^2+^ atom and a reference amino acid was measured over time.

### Phenotyping methods and analysis

Clinical information for all individuals with *de novo SYT1* variants was collated from clinical documentation and parent-report questionnaire (online or by post) using a standard template (for individual clinical histories see Table S3). The study-specific *Medical History Interview* gathered information about perinatal history, infant and child health, neurological symptoms and developmental milestones. The *Vineland Adaptive Behaviour Scales* is a standardised assessment of everyday adaptive functioning commonly used to support evaluation of neurodevelopmental disorders. Within the sample, either second edition^15^ (ID control n=34) or third edition^16^ (SYT1 n=14; ID control=17) of the Vineland were administered. The *Developmental Behaviour Checklist 2*^*17*^ *(DBC-P*) assesses emotional and behavioural problems in individuals with ID and comprises five subscales (disruptive/antisocial, self-absorbed, communication disturbance, anxiety and social relating). The *Social Responsiveness Scale*^*18*^ (*SRS*) is a standardised questionnaire enquiring about the presence and severity of social impairments (social motivation, social awareness, social cognition, social communication, and restricted interests and repetitive behaviour). The distributions of all outcome measures were examined for normality prior to parametric or non-parametric analyses as appropriate.

## Results

### 1. *SYT1* Variants

Overall, 15 *de novo* variants in *SYT1* were identified in 22 individuals (Figure 1a; 5 missense variants in 11 individuals have previously been described^8,9^). No alternative candidate variant potentially explaining neurodevelopmental presentation was identified in any case. ACMG criteria classified the variants as pathogenic (n=6), likely pathogenic (n=4) or uncertain (n=5; Table 1). The 11 newly-reported cases comprise one in-frame insertion (Lys367dup) and 10 missense variants (8 at novel loci, one at the previously reported Met303 locus, and one recurrent Ile368Thr variant). All missense variants are at highly evolutionarily conserved residues from human to invertebrates (COBALT; Table S4). While missense variation is not constrained across *SYT1* overall (observed to expected ratio = 0.47; gnomAD v2.1.1), the C2A and C2B domains lie within a region demonstrating significant missense constraint, with an observed to expected ratio of 0.24 (chi-square 48.87; ExAC).

**Table 1:** Assessment of SYT1 variant pathogenicity.

| Nucleotide and amino acid change | N in this paper | gnomAD v2.1.1 | SIFT | Polyphen-2 | M-CAP | Published functional evidence | ACMG classification | Predicted molecular impact |
| --- | --- | --- | --- | --- | --- | --- | --- | --- |
| c.476T>G<br>p.Leu159Arg (L159R) | 1 | 2 synonymous changes | Damaging (0) | Probably damaging (1.000) | Possibly pathogenic (0.085) | N | PM2, PM6, PP3<br>**VUS** | Structural perturbations in C2A |
| c.587C>A<br>p.Thr196Lys (T196K) | 1 | 2 synonymous changes (in controls) | Damaging (0) | Probably damaging (0.998) | Possibly pathogenic (0.057) | N | PM2, PM6, PP3<br>**VUS** | Structural perturbation in β-4 of C2A |
| c.625G>A<br>p.Glu209Lys (E209K) | 1 | Nil | Damaging (0) | Probably damaging (0.997) | Possibly pathogenic (0.029) | N | PM2, PM6, PP3<br>**VUS** | Removes H-bonds with Lys197 (between β-4 and β-5) |
| c.655G>C<br>p.Glu219Gln (E219Q) | 1 | Nil | Damaging (0) | Probably damaging (0.984) | Possibly pathogenic (0.039) | N | PM2, PP3, PM6<br>**VUS** | Neutralises surface negative charge and removes salt bridge with Lys223 |
| c.908T>A<br>p.Met303Lys (M303K) | 1 | 1 frameshift p.Met303TrpfsTer11 (in a neuro case) | Damaging (0) | Benign (0.329) | Possibly pathogenic (0.055) | $\Upsilon^9$ | PS3, PM1, PM2, PM6, PP3<br>**Pathogenic** | Structural perturbations in C2B |
| c.907A>G<br>p.Met303Val (M303V) | 1 | 1 frameshift p.Met303TrpfsTer11 (in a neuro case) | Tolerated (0.12) | Benign (0.322) | Possibly pathogenic (0.058) | N | PM1, PM2, PM5 (supporting), PM6<br>**Likely pathogenic** | Structural perturbations in $\text{Ca}^{2+}$-binding loop 1 of C2B |
| c.911A>G<br>p.Asp304Gly (D304G) | 1 | Nil | Damaging (0) | Possibly damaging (0.701) | Possibly pathogenic (0.229) | $\Upsilon^{9,10}$ | PS3, PM1, PM2, PM6, PP3<br>**Pathogenic** | Impaired $\text{Ca}^{2+}$-binding of C2B |
| c.925T>C<br>p.Ser309Pro (S309P) | 1 | Nil | Damaging (0) | Probably damaging (0.999) | Possibly pathogenic (0.130) | N | PM1, PM2, PM6, PP3<br>**Pathogenic** | Loss of H-bonds in C2B $\text{Ca}^{2+}$-binding loop 1 |
| c.1022A>G<br>p.Asn341Ser (N341S) | 1 | Nil | Damaging (0) | Probably damaging (0.992) | Possibly pathogenic (0.239) | N | PM2, PM6, PP3<br>**VUS** | May perturb interaction with SNAP-25 |
| c.1094A>G<br>p.Tyr365Cys<br>(Y365C) | 1 | Nil | Damaging<br>(0) | Probably<br>damaging<br>(0.993) | Possibly<br>pathogenic<br>(0.144) | N | PM1, PM2, PM6,<br>PP3<br>**Likely pathogenic** | Structural perturbations in<br>Ca<sup>2+</sup>-binding loop 1 of C2B |
| c.1098C>A<br>c.1098C>G<br>p.Asp366Glu<br>(D366E) | 3 | Nil | Damaging<br>(0) | Probably<br>damaging<br>(0.997) | Possibly<br>pathogenic<br>(0.061) | Y<sup>9,10</sup> | PS3, PM1, PM2,<br>PM6, PP3<br>**Pathogenic** | May alter Ca<sup>2+</sup> interaction |
| c.1100_1102dup<br>p.Lys367dup<br>(K367dup) | 1 | Nil | - | - | - | N | PM1, PM2, PM4<br>(supporting), PM6<br>**Likely pathogenic** | Add +ve charge and structural<br>perturbations to Ca<sup>2+</sup>-binding<br>loop 3 of C2B. May alter<br>membrane interaction. |
| c.1103T>C<br>p.Ile368Thr<br>(I368T) | 5 | Nil | Damaging<br>(0) | Benign<br>(0.186) | Possibly<br>pathogenic<br>(0.070) | Y<sup>8,9,10</sup> | PS3, PM1, PM2,<br>PM6, PP3<br>**Pathogenic** | Inhibited membrane<br>penetration |
| c.1106G>A<br>p.Gly369Asp<br>(G369D) | 1 | Nil | Damaging<br>(0) | Probably<br>damaging<br>(0.990) | Possibly<br>pathogenic<br>(0.077) | N | PM1, PM2, PM6,<br>PP3<br>**Likely pathogenic** | Add -ve charge to Ca<sup>2+</sup>-binding<br>loop 3 of C2B. May alter<br>membrane interaction. |
| c.1113C>G<br>p.Asn371Lys<br>(N371K) | 2 | 49 x synonymous<br>change (19 in<br>controls) | Tolerated<br>(1) | Probably<br>damaging<br>(0.999) | Possibly<br>pathogenic<br>(0.131) | Y<sup>9</sup> | PS3, PM1, PM2,<br>PM6, PP3<br>**Pathogenic** | May perturb structure of Ca<sup>2+</sup>-<br>binding loop 3 of C2B |

**Figure 1.**
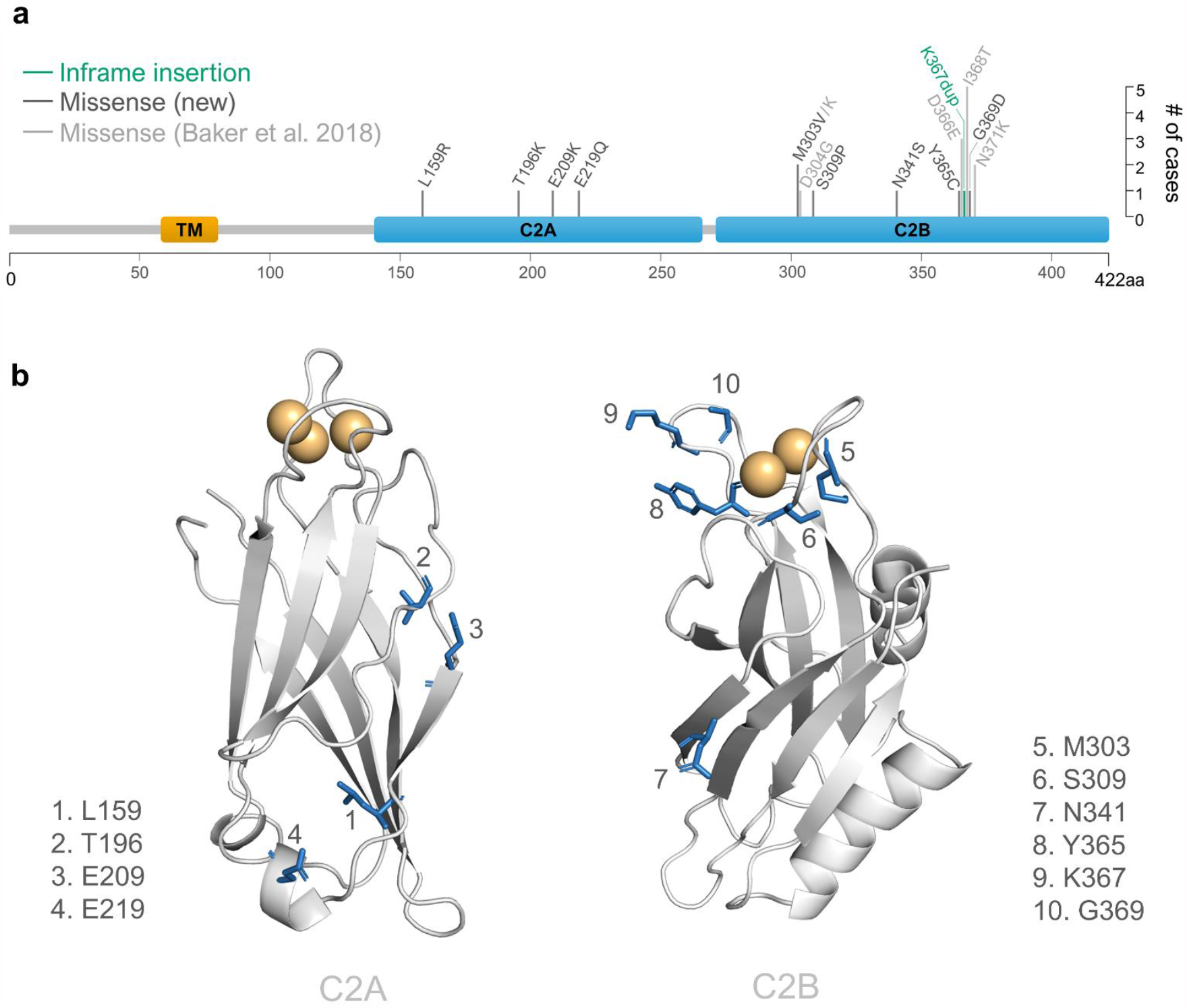
Location of newly-identified SYT1 variants. (**a**) Positions and types of newly-identified (dark grey) and previously described (light grey) variants are indicated on the domain structure of the *SYT1* gene. Length of the vertical line at each residue reflects the number of individual cases included in this study that harbour variants at that locus. TM: transmembrane domain. (**b**) Locations of newly-identified SYT1 variants are highlighted in 3D structures of the C2A (left; PDB: 1BYN) and C2B (right, PDB: 1K5W) domains with residues impacted by variants shown in blue as stick representation and numerically labelled. Ca^2+^ ions are represented as orange spheres.

### 2. Molecular Impacts of SYT1 Variants

To further investigate the pathogenic potential of SYT1 variants, we carried out molecular dynamics simulations of the mutated protein domains and searched for known roles of affected residues. For all newly-identified variants, the distances of Ca^2+^ ions from a reference amino acid in the Ca^2+^-binding pocket were comparable to the WT simulations (Figure S1), indicating that disturbed Ca^2+^ retention is unlikely to be a major pathogenic mechanism. In contrast, simulations of the previously reported Asp304Gly variant recapitulated defects in the retention of Ca^2+^ (Figure S1)^9^. Although no variant caused major structural changes to the C2 domain (Figure S2), many mutants altered the mobility of discrete regions of the domains (Figures 2, S3 and S4), or would be expected to impact intra- or intermolecular interactions (Figure 2, Table S5). These results suggest that disease-associated variants are likely to impact the structure or function of SYT1 through diverse molecular mechanisms. Literature-informed predictions of molecular impacts and molecular dynamics simulation results are described below for each newly-reported variant.

**Figure 2.**
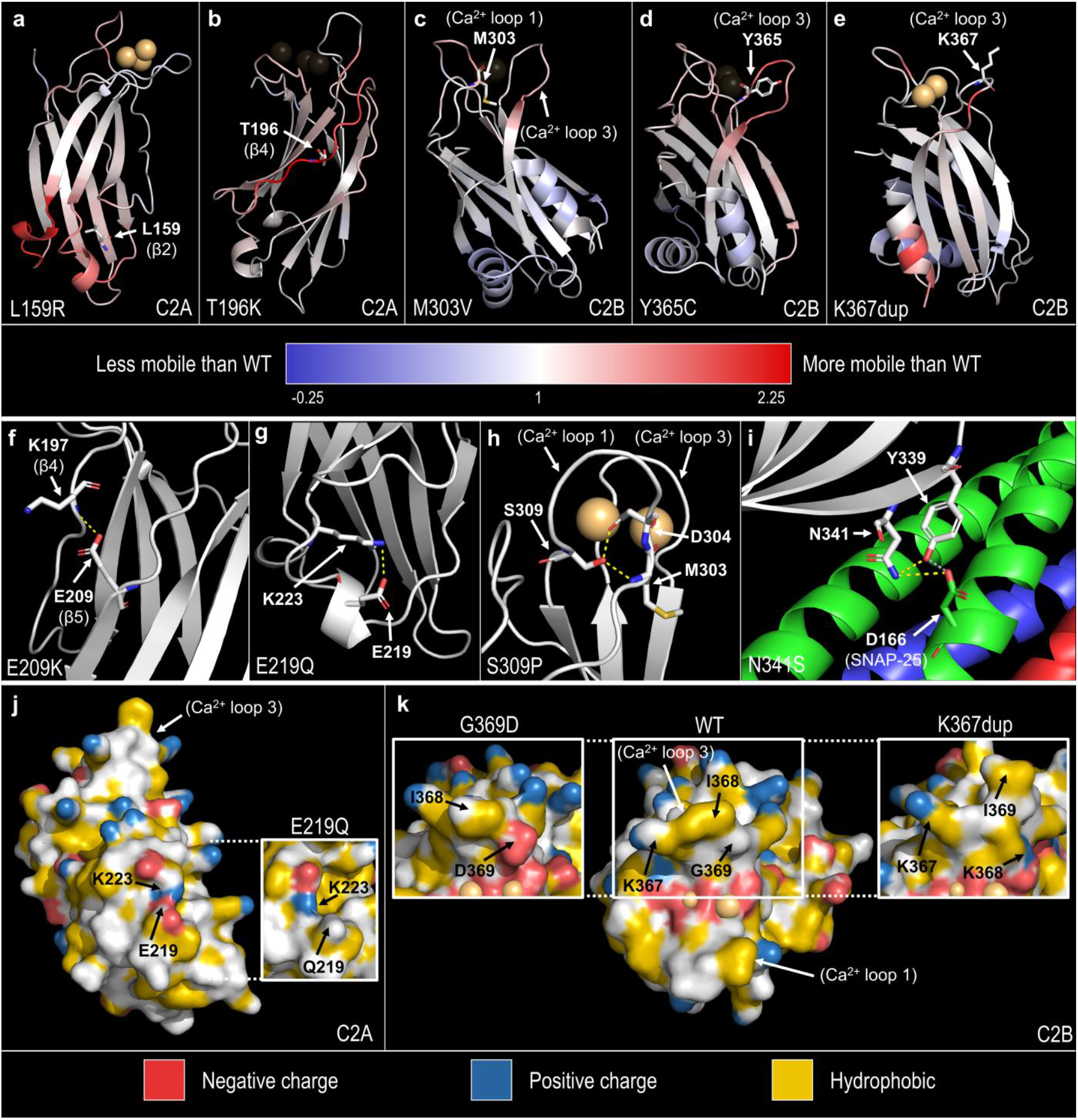
Predicted molecular impacts of newly-identified SYT1 variants. Variants are expected to alter regional mobility of the domain (**a-e**), perturb intra- or intermolecular interactions (**f-i**), or alter the surface charge of the protein (**j**,**k**). (**a-e**) Cartoon ribbon representations of WT C2A and C2B domains where blue-white-red spectrum colouring indicates the change in mobility of each residue between WT and mutant simulations. Mobility change was calculated as average mutant RMSF (Å)/average WT RMSF (Å) for each residue and thresholded at -0.25 (blue) and 2.25 (red) for illustration with 1 denoting RMSF equal to WT (white). Residues mutated in SYT1 cases are shown in stick representation and labelled. Either Ca^2+^-bound structures (**a**,**e**) or Ca^2+^-free structures (**b-d**) are shown to display the greatest impact on RMSF. (**f-h**) Intramolecular interactions that were abolished in simulations of mutant domains are shown as yellow dotted lines (see Table S5 for details). (**f**) C2A domain showing the Glu209-Thr196 hydrogen bond lost in Glu209Lys. (**g**) C2A domain showing the Glu219-Lys223 salt-bridge lost in Glu219Gln. (**h**) C2B domain showing Ser309-Met303 and Ser309-Asp304 hydrogen bonds lost in Ser309Pro. (**i**) Primary interface of the SYT1-SNARE complex (PDB: 5CCH; white: SYT1, green: SNAP-25, red: synaptobrevin-2, blue: Syntaxin-1A) highlighting the intramolecular (Asn341-Tyr339 in SYT1) and intermolecular (Asn341^SYT1^-Asp166^SNAP-25^) hydrogen bonds involving Asn341 in yellow. (**j**,**k**) YRB representations^37^ of Ca^2+^-bound WT and mutant C2A or C2B domains (at last frame of simulation) that show surface charge and hydrophobicity (red: negative charge, blue: positive charge, yellow: hydrophobic). (**j**) WT and Glu219Gln (inset) C2A domains. (**k**) WT, Gly369Asp (left inset) and Lys367dup (right inset) C2B domains. Ca^2+^ ions are represented as orange spheres.

All four C2A substitutions occur in regions of undetermined function. Leu159, lying in β-2, faces the hydrophobic interior of the domain. The side chain of the introduced charged arginine (Leu159Arg) inserts between the β-sheets and disrupts the stability of the region, with elevated RMSF in multiple regions, particularly across the bottom (non-Ca^2+^-binding) loops of the C2A domain (Figures 2a and S3). Two substitutions, Thr196Lys and Glu209Lys, are in close proximity to each other in opposing β-strands (β-4 and β-5, respectively) on the edge of the C2A β-sandwich (Figure 1b). Thr196 is structurally important with its side chain buried between β-3 and β-4, inducing a distortion in the β-sheet structure^19^. Thr196Lys has substantially increased RMSF over residues Lys190 to Lys201 (Figure 2b and S3), indicating localised instability possibly arising from impaired anchoring of β-4 to the rest of the β-sheet. Glu209 forms hydrogen bonds with Lys197 in β-4 of the opposing β-sheet that are removed by lysine substitution (Figure 2f, Table S5). However, there is no observable alteration to RMSF of the Glu209Lys mutant (Figure S3). Glu219 is situated in a short alpha-helix distal to the Ca^2+^- binding loops; its side chain points away from the protein and could possibly participate in intermolecular interactions. Glu219Gln neutralises the negative charge at that site and removes a salt bridge between Glu219-Lys223 (Figures 2g and 2j, Table S5), but does not result in an appreciable effect on regional RMSF (Figure S3).

Similar to previously reported cases^9^, all newly-identified C2B missense variants map to the region surrounding the Ca^2+^-binding pocket (Met303Val and Ser309Pro in loop 1; Tyr365Cys, Lys367dup, and Gly369Asp in loop 3) with one notable exception, Asn341Ser, located in a β-strand (Figure 1b).

Met303 anchors Ca^2+^-binding loop 1 and replacement with valine is predicted to render the loop more flexible, with increased RMSF across residues 301-306 in Ca^2+^-free Met303Val simulations (Figures 2c and S4). Ser309Pro would abolish transient hydrogen bonds between Ser309-Met303 and Ser309-Asp304 that are normally present in the Ca^2+^-bound WT C2B domain (Figure 2h, Table S5), but no change to the RMSF of Ca^2+^-binding loop 1 was observed in Ser309Pro simulations (Figure S4). In the Ca^2+^-bound simulations, both Met303Val and Ser309Pro increased the mobility of the distal arginine apex (Arg399, Arg400) at the opposite end of the C2B domain (Figure S4).

No structural alterations were detected in simulations of the Asn341Ser mutant (Figure S4), but importantly Asn341Ser may perturb the interaction between SYT1 and the SNARE complex. Asn341 faces outwards on β-5 and is proximate to the primary binding interface between SYT1 C2B and SNAP-25^20^. Crystal structures of the SYT1-SNARE complex (PDB: 5CCH, 5CCG and 5KJ7)^20^ show that Asn341 may interact directly with Asp166 of SNAP-25 or form a hydrogen bond with the neighboring Tyr339 in SYT1, which binds Asp166 of SNAP-25 (Figure 2i). Interestingly, two variants at Asp166 have been identified in individuals with *SNAP25* developmental and epileptic encephalopathy^21^.

Tyr365 stabilises Ca^2+^-binding loop 3 and the mobility of this loop is increased in Tyr365Cys mutant simulations (Figures 2d and S4). Furthermore, Asp366 flips out of the Ca^2+^-binding pocket, and Lys367 twists to impinge on the Ca^2+^-binding pocket in Ca^2+^-free Tyr365Cys simulations. The positive charge at Lys367 is important for phospholipid binding^22-24^. The Lys367dup variant introduces an additional positively charged lysine to Ca^2+^-binding loop 3 (Figure 2k), and thus could potentially increase attraction between the tip of loop 3 and anionic phospholipids. Additionally, simulations showed increased flexibility in loop 3 (Figures 2e and S4) and transient interactions between the inserted lysine and Ca^2+^-coordinating residues Asp366, Asp373, Asp310 and Asp304. Gly369Asp introduces an additional negative charge to the Ca^2+^-binding pocket (Figure 2k), more specifically the membrane-penetrating tip of loop 3, which could be expected to repel the anionic plasma membrane.

### 3. Clinical histories

In keeping with previous findings, the most common features within the SYT1 group were developmental delay, ophthalmic problems and abnormal EEG (cohort summary in Table 2; individual data in Table S3). There is a wider range of severity of neurodevelopmental impairments than previously reported – approximately one third of cases presented with mild or moderate delay to motor and communication milestones, whereas almost all previously reported cases were severely delayed^8,9^. Other common features were sleep disorder, feeding difficulties, gastrointestinal reflux, and finger-chewing or other self-injury, each affecting around two thirds of cases.

**Table 2.** Clinical phenotypes summary.

| *Clinical feature* | *Data available / n* | *Frequency of feature / n* | *Frequency of feature / %* | *Subtype (n)* |
| --- | --- | --- | --- | --- |
| *Delayed communication milestones* | 22 | 21 | 95 | Mild = using words and phrases (5), Moderate = using single words only (2), Severe = not using any words (6), Unable to classify as under age 5 years or insufficient information (8) |
| *Abnormal EEG* | 18 | 17 | 94 | Ictal features (8), intermittent low frequency oscillations (8), abnormal background activity unspecified or generalised slowing (6) |
| *Ophthalmic* | 22 | 20 | 91 | Strabismus / esotropia (12), nystagmus (6), hypermetropia (3), visual impairment unspecified (3) |
| *Infantile hypotonia* | 22 | 19 | 86 |  |
| *Delayed motor milestones* | 22 | 18 | 81 | Mild = walked by 3 years (4), Moderate = walked by 5 years (3), Severe = walked after 5 years or non-ambulatory over the age of 5 (6), unable to classify as non-ambulatory under age 5 years (5) |
| *Movement disorder* | 21 | 14 | 66 | Dystonia (7), chorea (7), dyskinesia (1), ataxia (5), myoclonus (3), tremor (2) stereotypies (6) |
| *Sleep disorder* | 19 | 12 | 63 | Commonly hypersomnia during infancy then difficulties initiating and maintaining sleep |
| *Gastrointestinal* | 22 | 13 | 59 | Feeding difficulties (8), gastroesophageal reflux (6), drooling (2), constipation (3), chronic diarrhoea (1), urinary retention (2), pancreatitis with pseudocysts (1) |
| *Finger chewing or other self-injury* | 22 | 13 | 59 | Finger biting or chewing (9), head banging (3), skin picking (1), other or unspecified (1) |
| *Musculoskeletal* | 22 | 8 | 36 | Torticollis (1), joint hypermobility (2), talipes (2), pes planovalgus (1), progressive contractures (1), scoliosis (1) |
| *Abnormal MRI* | 15 | 5 | 33 | Mild diffuse progressive volume loss (1), delayed myelination (2), mild periventricular WM nodular abnormality (2) |
| *Airways / respiratory / cardiology* | 22 | 6 | 27 | Sleep apnoea (4), laryngomalacia (1), hyperventilation with cyanosis (1), autonomic dysfunction with hypotension (1) |
| *Minor congenital abnormalities* | 22 | 5 | 22 | Undescended testicle (1) Atrial septal defect (1) Dermoid cyst (2) Unilateral syndactyly (1) |
| *Epilepsy diagnosis* | 22 | 4 | 18 | Absence seizures (3), tonic-clonic seizures (1), infantile spasms (1) |
| *Perinatal problems* | 22 | 3 | 13 | Mild prematurity (1) Neonatal resuscitation (1) Meconium aspiration (1) |

Movement disorder is a feature in two thirds of the cohort, a higher frequency than previously reported^9^. The types and severities of involuntary movements are variable and include ataxia, tremor and myoclonus, as well as dystonia, chorea and complex hyperkinetic movement disorders in more severely affected cases.

In contrast to our previous case series with no cases of epilepsy^9^, four individuals in the current group have received an epilepsy diagnosis. Reported seizure phenotypes include absences (n=3), tonic-clonic seizures (n=1) and infantile spasms (n=1). Additional cases are required to clarify the association between *SYT1* and epilepsy risk.

EEG abnormalities remain very common across the cohort, encompassing low frequency background oscillations as previously reported, but also ictal features in individuals with and without overt seizures.

Although the majority of individuals within the group are young children, it is possible to make some preliminary comments on the longer-term trajectory of the condition based on 9 individuals currently over the age of 10 years. We note the potential for long-term positive progress in motor development, with two individuals learning to walk after the age of 5 years. However, two individuals have developed movement disorder symptoms during late childhood, accompanied by relative decline in adaptive skills. We also observe that social and emotional difficulties may emerge with time, with some older children and adolescents experiencing obsessions, anxieties and mood disturbance.

### 4. Behavioural phenotyping and comparison to ID control group

Vineland Adaptive Behaviour Composite (ABC) scores within the SYT1 group ranged from 20 to 74. Four participants scored in the borderline or mild ID range, three in the moderate ID range, and seven in the severe or profound ID range (see Figure 3a for breakdown of subscale scores for each individual). Inspecting Vineland subscale scores within the SYT1 group, we observed that communication ability is more severely impaired on average than motor abilities, socialisation and daily living skills. In contrast, motor ability is a relative strength within the ID comparison group. To establish whether these differences reflect a consistent and discriminating profile of adaptive functions within the SYT1 group we carried out General Linear Model analysis (within-subjects factor: Vineland Subscale; between subjects factors: Group). This highlighted significant interaction between Group and Subscale (*F*=4.54, *df* 2.33 Greenhouse Geisser corrected, *p*=0.01; reducing to p=0.09 after co-varying for Vineland edition). Post-hoc non-parametric analyses indicated significantly lower scores for the SYT1 group in Motor ability (*p*=0.036) and Communication (*p*=0.019) but not Socialisation (*p*=0.07) or Daily Living skills (*p=*0.28) (Figure 3b).

**Figure 3.**
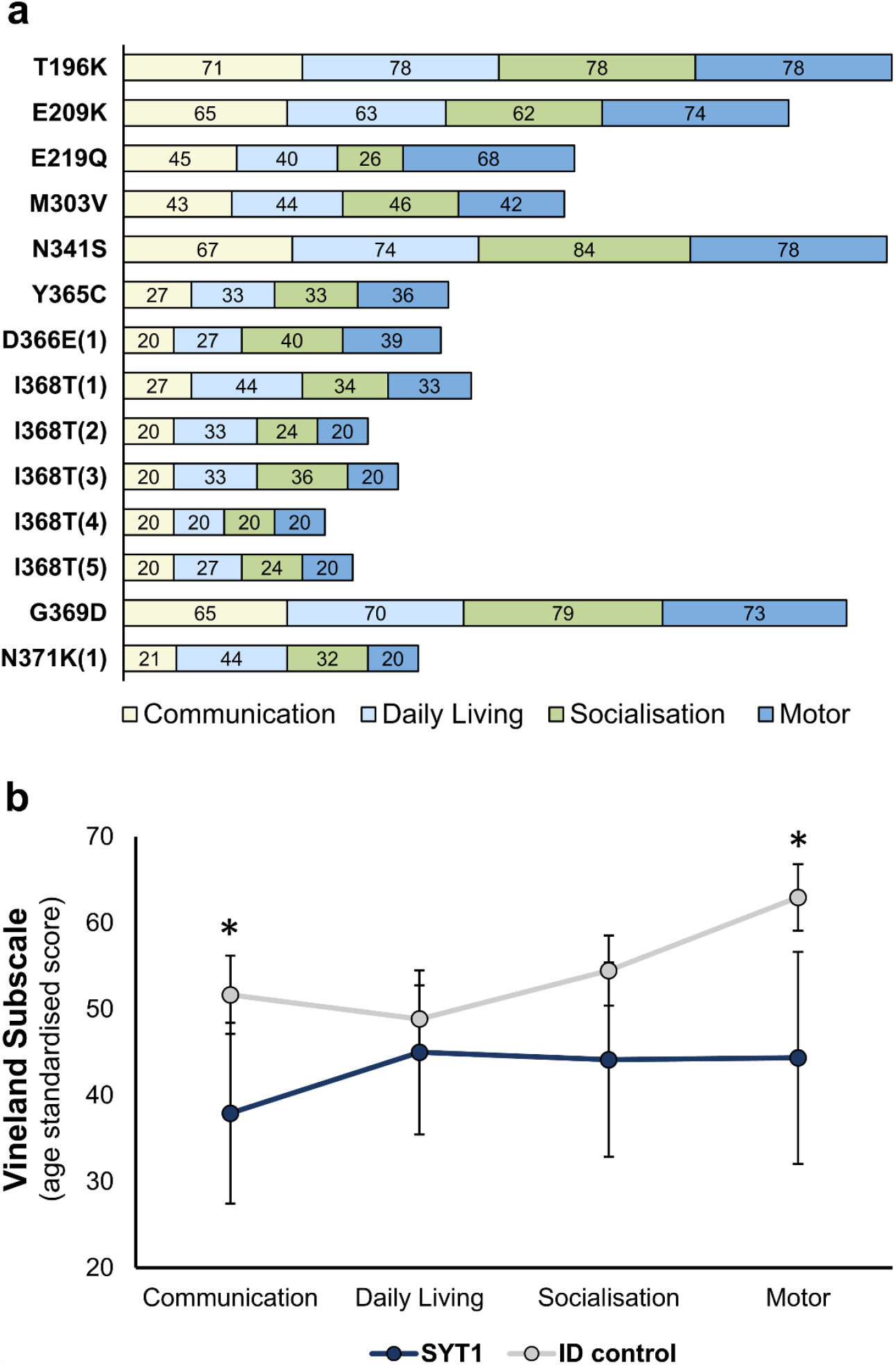
Adaptive behaviour profiles of SYT1 and comparison groups. (**a**) Standardised scores for Vineland subdomains are shown for each SYT1 individual assessed. Case numbers corresponding to individuals listed in Table S3 are indicated in parentheses for recurrent variants. (**b**) Comparisons of group averages of Vineland subdomains for SYT1 (n=14) and ID control (n=51) groups (uncorrected for Vineland form). Error bars represent 95% confidence intervals. General linear model analysis revealed significant differences between groups in Communication and Motor subdomains (\**p*<0.05).

We next explored DBC-P and SRS-2 scores, assessing emotional or behavioural problems and autism-related social impairment respectively. No significant differences emerged between SYT1 and ID comparison groups for total scores on either measure, and GLM analyses identified no significant Group x Subscale interaction for either measure (Figures S5 and S6).

In our previous description of *SYT1*-associated phenotypes^4^, based on clinical reports only, we noted that a high proportion of diagnosed individuals displayed motor stereotypies, unpredictable mood switches, and episodes of agitation. To explore whether these features are increased in the expanded SYT1-diagnosed population, beyond expectation for ID, five relevant items reflecting these symptoms were selected from the DBC-P (6. Bangs head; 10. Chews or mouths body parts; 33. Hits or bites self;

47. Mood changes rapidly for no reason; 60. Has repeated movements of hands, body or head). These items appear across two DBC-P subscales (disruptive/antisocial, self-absorbed). Raw scores (0-2) for each item were summed for each participant. Univariate analysis, co-varying for age, identified significantly higher scores for these selected DBC items within the SYT1 group (SYT1 group *M*= 5.57, *SD*= 2.95, range 0-9; ID comparison group *M*=3.67, *SD*=2.98, range 0-10; *F*=5.95, *df* 1, *p*=0.02). Age-adjusted summed scores showed a strong negative correlation with Vineland ABC within the SYT1 group (Spearman’s rho -0.73, *p*=0.003), but not the comparison group (Spearman’s rho -0.18, *p*=0.26; Fisher’s test^25^ *z*=-2.19, *p*=0.01) (Figure S7). In summary, self-injury, mood instability and repetitive movements are elevated within the SYT1 group, with a strong relationship between these features and global adaptive impairment.

## Discussion

This work builds on the previous identification and characterisation of *SYT1*-associated neurodevelopmental disorder to broaden the range of potentially pathogenic variants for clinical laboratory reporting. We detail the expansion of the genotypic and phenotypic spectrum of this syndrome with the objective of increasing the diagnostic efficiency of this ultra-rare disorder as well as improving prognosis estimation, patient management, and information available for families and clinicians after diagnosis.

### 1. Broadening the genetic landscape

We describe four novel variants in the SYT1 C2A domain as well as additional C2B loci, revealing that neurodevelopmental disorder-associated variants are found in the two functional C2 domains of SYT1. Whilst C2A variants remain of uncertain significance according to ACMG criteria, and functional evidence is required to substantiate pathogenicity, molecular dynamics simulations and existing literature are supportive of likely functional impact. Novel variants may exert dominant-negative effects, as seen for previously identified mutants^8-10^, but we cannot preclude haploinsufficiency as a possible pathogenic mechanism. Ca^2+^-binding loops 1 and 3 of the C2B domain appear to be highly sensitive to mutation as disease-associated missense variants cluster in these regions in both *SYT1* and the homologous *SYT2*^26,27^, and no missense *SYT1* variants in these loops are recorded in gnomAD v2.1.1. Therefore, any *de novo* missense variant in the C2B Ca^2+^-binding loops should be investigated for possible pathogenicity and variants in other highly conserved residues of the SYT1 C2 domains should also be considered.

### 2. Broadening the phenotypic spectrum

The major clinical features associated with *SYT1* variants in this larger cohort are broadly in keeping with those reported in the previous case series^9^. *SYT1*-associated neurodevelopmental disorder presents with individually non-specific features, but may be suspected when neonatal hypotonia, developmental delay, ophthalmic problems, movement disorders and EEG abnormalities are present in any combination. While some of the newly-identified cases present with profound developmental delay and involuntary movement disorders, in line with the presentation of initial cases, others show milder neurodevelopmental difficulties thereby widening the range of clinical severity. For these less severe cases, it is more difficult to distinguish a discriminating phenotypic signature to aid in variant interpretation and diagnostic confirmation. Given the young age of some individuals within the cohort and uncertainty of the developmental trajectory of this disorder, a cautious prognosis is warranted. We note that amongst some older diagnosed cases, later-onset movement disorder and decline in adaptive function and emotional wellbeing have been observed. Longitudinal sampling of individuals with *SYT1* variants will facilitate mapping of the developmental progression of this disorder and improve prognosis estimations.

Questionnaire data revealed that all domains of adaptive function are impaired within the SYT1 cohort, and highlighted disproportionate impact on motor and communication function compared to a comparison ID group (although note that data could not be collected for 7 SYT1 cases due to language barriers or young age). Moreover, features of rapid mood change and motor stereotypies (specifically hand biting) are prominent, in line with previous observations^9^. Improved understanding of the cell-type specific dependence on SYT1 for efficient neurotransmitter release will aid symptom explanation at the circuit and large-scale network levels. Future investigation will involve additional parent/carer-report questionnaires to further probe neurological and behavioural domains impacted by *SYT1* mutations, such as visual behaviour, movement disorder symptoms, repetitive behaviours and hyperactivity.

With expansion of the cohort, *SYT1*-associated neurodevelopmental disorder continues to demonstrate intersecting clinical features that are common amongst synaptic vesicle cycling disorders^3^. However, seizures are a significant feature of disorders associated with mutation of other core synaptic vesicle fusion machinery including *VAMP2, STX1B, SNAP25, STXBP1, CPLX1* and other accessory proteins^28,29^. In notable contrast, while four SYT1 cases have reported seizures, epilepsy is evidently not a prevalent or prominent feature in this disorder. While EEG abnormalities were frequent within the SYT1 group, the electrophysiological features observed were variable and there were inconsistencies in reporting methods, provision of recordings for review, patient age and conditions of recordings. Standardised and systematic EEG data collection and analysis will be needed to confirm common electrophysiological characteristics and inform understanding of the neurophysiological origins of symptoms. Furthermore, systematic behavioural characterisation of other disorders of fusion, and synaptic vesicle cycling disorders more broadly, will allow more detailed comparison of these mechanistically-related syndromes and interrogation of distinctive characteristics of this group of disorders.

### 3. Diversity of molecular mechanisms

Molecular dynamics simulations and SYT1 structure-function relationships sourced from the literature were used to carefully consider variants in the context of the 3D protein structure. Newly-identified variants are predicted to have nuanced effects on the local stability of discrete protein regions (including Ca^2+^-binding loops or β-sheets), penetration of the phospholipid membrane, or interaction with SNARE proteins, rather than grossly destabilise the C2 domain or substantially impair Ca^2+^ binding. Such perturbations could be expected to disrupt SYT1 function and synaptic transmission. Some disorder-associated variants, particularly those within the C2A domain, provide the first indication of the importance of previously unrecognised residues and regions of SYT1. It should be noted that while molecular dynamics simulations provide atomic-level predictions of variant impact on protein structure and Ca^2+^ retention, these simulations are limited in length and unable to reveal impacts on protein-protein or protein-lipid interactions or consequences on neurotransmission. Functional studies at the molecular, cellular and circuit levels will provide further evidence for pathogenicity and insight into the specific mechanisms underlying neurodevelopmental impairments.

### 4. Genotype-phenotype links

We inspected the current datasets for evidence that the specific *SYT1* variant may contribute to clinical features and severity. Questionnaire data was available for five cases with the recurrent Ile368Thr variant, which revealed that the neurological and behavioural phenotype was highly consistent between these individuals (Figure 3, Table S3). Although questionnaire data was not available for other recurrent variants, clinical reports of recurrent locus Met303 and variants Asp366Glu and Asn371Lys show similar consistency in phenotype severity (Table S3). This raises the possibility of a relationship between diversity of molecular mechanisms and phenotypic variation. No obvious patterns emerged between the clinical phenotype and either the nature of the amino acid substitution or the conservation of the affected residue across synaptotagmin isoforms.

We subsequently asked whether there are consistent differences between C2A and C2B domain variants in global severity of impairments. While acknowledging that cases of C2A variants are limited, it is notable that all four of these individuals present with mild or moderate constraints to adaptive function and absence of early-onset movement disorder. Correspondingly, the variants linked to the most severe clinical phenotypes are all situated in the C2B domain. The prospect of differential clinical severity between the C2A and C2B domains is congruent with the current understanding that each C2 domain plays a distinct role. Specifically, the C2B domain is essential for Ca^2+^-dependent membrane binding and fast synchronous neurotransmitter release, while the C2A domain may be facilitatory and mediate other modes of release^30-34^. However, there is not an absolute divide in clinical severity between C2A and C2B domains, as not all C2B variants are severe with several cases of intermediate severity and two notably milder novel variants (Asn341Ser and Gly369Asp). It is also important to note that no identified C2A variant is located around the Ca^2+^-binding loops (in contrast to most C2B variants), which may also account for this differential presentation. Most C2A variants fall within β-strands and could have diffuse structural impacts that differ from specific disruption of the Ca^2+^- binding loops. Indeed, the only C2B variant outside the Ca^2+^-binding loops (Asn341Ser) is associated with comparatively milder features. However, mildly presenting Gly369Asp is situated in a C2B Ca^2+^- binding loop, arguing that location within the domain does not completely explain clinical severity. Identification of additional variants and detailed phenotyping of recurrent variants will assist in clarifying genotype-phenotype relationships. Further, the impacts of variants on all functions of SYT1, not only evoked exocytosis but also suppression of spontaneous and asynchronous release and modulation of endocytosis^35,36^, need to be investigated to fully appreciate similarities and differences in molecular and cellular impacts. Integration of genetic, cellular, neural systems and cognitive investigations will enable a thorough understanding of *SYT1*-associated neurodevelopmental disorder, with the prospect of precision medicine targeting each individual’s symptoms and underlying mechanisms.

## Data Availability

Reporting of SYT1 variants in open access repositories is listed in Table S3. Molecular dynamics simulation data not included in the Supplementary Material are available upon request to the corresponding author.

## Data Availability

Reporting of *SYT1* variants in open access repositories is listed in Table S3. Molecular dynamics simulation data not included in the Supplementary Material are available upon request to the corresponding author.

## Acknowledgements

We appreciate the important and generous contributions of each individual with a *SYT1* variant, their families and carers. We appreciate the roles of each clinician and laboratory scientist involved in the diagnostic pathway of each case. Further, we appreciate the involvement of individuals in the comparison ID group and their families and carers. We acknowledge the *NIHR UK Rare Genetic Disease Research Consortium* for assistance with recruitment to the study. HM is supported by an Australian Government Research Training Program Scholarship. SLG and HM are supported by Grant 2003710 from The National Health and Medical Research Council (NHMRC) of Australia and a Florey Fellowship. The Florey Institute of Neuroscience and Mental Health acknowledges the strong support from the Victorian Government and in particular the funding from the Operational Infrastructure Support Grant. This project was supported by funding to KB from the UK Medical Research Council (G101400), the Baily Thomas Charitable Trust, and Great Ormond Street Hospital Children’s Charity. FB and JJZ were supported by the Indiana University Precision Health Initiate. Molecular dynamics simulations were supported in part by Lilly Endowment, Inc., through its support for the Indiana University Pervasive Technology Institute. We thank Dr. Trayder Thomas (University of Chicago) and Dr. Billy Williams-Noonan (RMIT University) for assistance with molecular dynamics simulations. The study makes use of data generated by the DECIPHER community. A full list of centres who contributed to the generation of the data is available from https://decipher.sanger.ac.uk and via email from. Funding for the DECIPHER project was provided by Wellcome Trust. Individuals who contributed to DECIPHER bear no responsibility for the interpretation of the data by the authors. We acknowledge the DDD study for identification of variants reported in this paper. The DDD study presents independent research commissioned by the Health Innovation Challenge Fund [grant number HICF-1009-003] a parallel funding partnership between the Wellcome Trust and the Department of Health, and the Wellcome Trust Sanger Institute [grant no. WT098051]. See Nature 2015; 519:223-8 or www.ddduk.org/access.html for full acknowledgement.

## Author Information

### Author Contributions

Conceptualization: K.B., H.M., S.L.G.; Formal Analysis: F.B. K.B., A.K.-T., A.J. Funding acquisition: K.B., S.L.G.; Investigation: F.B., H.M., A.K.-T., K.B., E.N.-C., A.J.; Resources: J.J.Z., P.C., S.J., M.L., C.F., J.T., F.E., M.W., T.K., R.P., R.B., R.C., L.L., M.A., J.v.d.S., M.N., M.K.; Visualization: H.M., A.K.-T., F.B.; Writing –original draft: H.M., K.B., F.B.; Writing – review & editing: H.M., K.B., S.L.G., D.J.S, all other authors reviewed and approved this manuscript.

## Ethics Declarations

This study was conducted within the “Phenotypes in Intellectual Disability” project, which received approval from Cambridge Central Research Ethics Committee (REC ref: IRAS 83633). Written informed consent was provided by each diagnosed individual’s parent or consultee prior to information sharing by referring clinicians, and questionnaire completion by parents or carers.

## Competing interests

The authors declare no conflict of interest.

## Supplementary Material

### Supplementary Methods

#### Evaluation of Variants

**gnomAD v2.1.1** (The Genome Aggregation Database) (https://gnomad.broadinstitute.org/) was searched to identify allele frequency (AF) of reported variants in control populations. To identify the evolutionary conservation of the amino acids in the protein sequence across 14 species from Human to *Caenorhabditis elegans*, we employed **COBALT** (Constraint-based Multiple Alignment Tool) (https://www.ncbi.nlm.nih.gov/tools/cobalt/cobalt.cgi). We deployed three *in-silico* predictive programmes, namely **SIFT version 5.2.2** (Sorting Intolerant From Tolerant) (http://sift.jcvi.org) to predict the effect of amino acid substitution on protein function, **PolyPhen-2** (Protein Analysis Through Evolutionary Relationship) (http://genetics.bwh.harvard.edu/pph2/) to predict deleteriousness of single nucleotide variants and insertions/deletions variants, **M-CAP** (Mendelian Clinically Applicable Pathogenicity) (http://bejerano.stanford.edu/mcap/) to combine previous pathogenicity scores (SIFT, PolyPhen) to increase the overall sensitivity. Evidence for pathogenicity and classification were carried out independently by two authors. Phenotypic similarity (supporting criterion PP4 in Richards et al. 2015^1^) was not applied, to avoid bias of our analysis of phenotypic spectrum.

#### Molecular Dynamics Simulations

MD simulations were carried out based on NMR structures of the synaptotagmin-1 (SYT1) C2A (PDB: 1BYN^2^) and C2B (PDB: 1K5W^3^) domains, or mutant variants thereof, either in the presence or absence of bound Ca^2+^ atoms. Single mutant homology models of SYT1 C2A comprising mutations Leu158Arg, Thr195Lys, Glu208Lys and Glu218Gln, and SYT1 C2B comprising mutations Met302Val, Asp303Gly, Asn340Ser, Tyr364Cys and Gly368Asp were generated using Swiss-PdbViewer v4.1.0, while the homology model containing the Lys366 duplication (K366dup) was generated using SWISS-MODEL^4^ (note that numbering used throughout this paper follows human sequence for simplicity (i.e. Leu159Arg, Thr196Lys, Glu209Lys and Glu219Gln, Met303Val, Asp304Gly, Asn341Ser, Tyr365Cys and Gly369Asp). The SYT1 C2A models were C- and N-terminally capped with Ace and NMe groups, respectively, to eliminate aberrant interactions with Ca3. Ca^2+^ atoms were removed from homology models prior to equilibration for simulations without Ca^2+^. All MD simulations were performed with the Gromacs molecular dynamics simulation package running version 2019.4 or 2020.3^5^ using the Amber ff99SB-ILDN forcefield^6^ under periodic boundary conditions and in a rhombic dodecahedron unit cell. Each system was solvated with simple point charge (SPC) water molecules^7^ and Na^+^ and Cl^−^ ions were added to neutralize the total charge of the systems at concentrations of 150 mM. The neighbour list, Coulomb and van der Waals interaction cut-offs were set to 1 nm and the particle mesh Ewald (PME) algorithm^8^ was utilised for long-range electrostatic interactions. All systems were first subjected to a 2000 step steepest descent energy minimization, alternatively completing when the maximum force on any atom has reached 1000 kJ mol^-1^ nm^-1^. A 1 ns NVT (constant Number of particles, Volume and Temperature) was then performed, heating the systems to 310 K using the V-rescale thermostat^9^ with position restraints on. This was followed by a 1 ns NPT (constant Number of particles, Pressure and Temperature) equilibration run performed with position restraints on using the V-rescale thermostat^9^ and the Berendsen barostat^10^. A third equilibration phase using randomised initial velocities, the V-rescale thermostat^11^ (310 K) and the Parrinello-Rahman barostat^12^ was performed for 10 ns using a 2 fs time step. Four individual production runs of ∼400ns were carried out for each system under the same conditions as the final equilibration phase but with position restraints switched off. We expected to observe nuances across four simulations that may not be observable in a single trajectory and, importantly, results for WT and Asp304Gly structures obtained with these four shorter simulations are in good agreement with previous results for single ∼1200ns trajectories based on the same starting models^13^. The MD trajectories were simplified by extracting every nanosecond and by removing all water molecules using the GROMACS trjconv utility. All simulations were carried out on the ‘Big Red 3’ system provided by the Indiana University Pervasive Technology Institute, Bloomington IN. Data analysis was carried out in VMD (v1.9.3) using the RMSF, RMSD and hydrogen bond (cut-off set to 3.5Å and 20°) plugins and Graphpad Prism (v9.0.0), and images of molecular snapshots were generated with PyMOL (v2.4.0). To measure Ca^2+^ retention, the distance between each Ca^2+^ atom and the C^γ^ of Asp231 of C2A or Asp364 of C2B was measured for all trajectories and plotted as a function of simulation time. Asp231 and Asp364 were used as reference amino acids as they are directly involved in electrostatic interactions with Ca1 and Ca2 in each C2 domain and are the most stable among Ca^2+^-interacting residues.

While Ca1 and Ca2 of both C2 domains remained stably bound in WT simulations (Figure S1), Ca3 occupancy of the C2A Ca^2+^-binding pocket was remarkably unstable with Ca3 dissociating from the WT domain at early timepoints in all four trajectories (Figure S8). Ca3 also dissociated from each mutated C2A domain (Leu159Arg, Thr196Lys, Glu209Lys, Glu219Gln) in at least one trajectory (Figure S8). Early Ca3 dissociation is reasonable given the low affinity of Ca3 (*K*_d_ of >>1mM)^14,15^, and it is unlikely that any of the mutations make a significant contribution towards stabilising Ca3 as a correlation with RMSF values of residues involved in Ca3 binding could not be established.

### Supplementary Data

**Supplementary Table 1.** Genetic diagnosis summary for the ID comparison group.

| Gene | n | % |
| --- | --- | --- |
| ARID1B | 11 | 21.6 |
| CASK | 2 | 3.9 |
| CTNNB1 | 1 | 2.0 |
| DDX3X | 15 | 29.4 |
| DLG3 | 2 | 3.9 |
| DYRK1A | 3 | 5.9 |
| EHMT1 | 5 | 9.8 |
| GRIN2A | 1 | 2.0 |
| KAT6B | 1 | 2.0 |
| PAK3 | 1 | 2.0 |
| SETD5 | 6 | 11.8 |
| SHANK3 | 2 | 3.9 |
| SMARCA2 | 1 | 2.0 |
| =51 |  |  |

**Supplementary Table 2.**
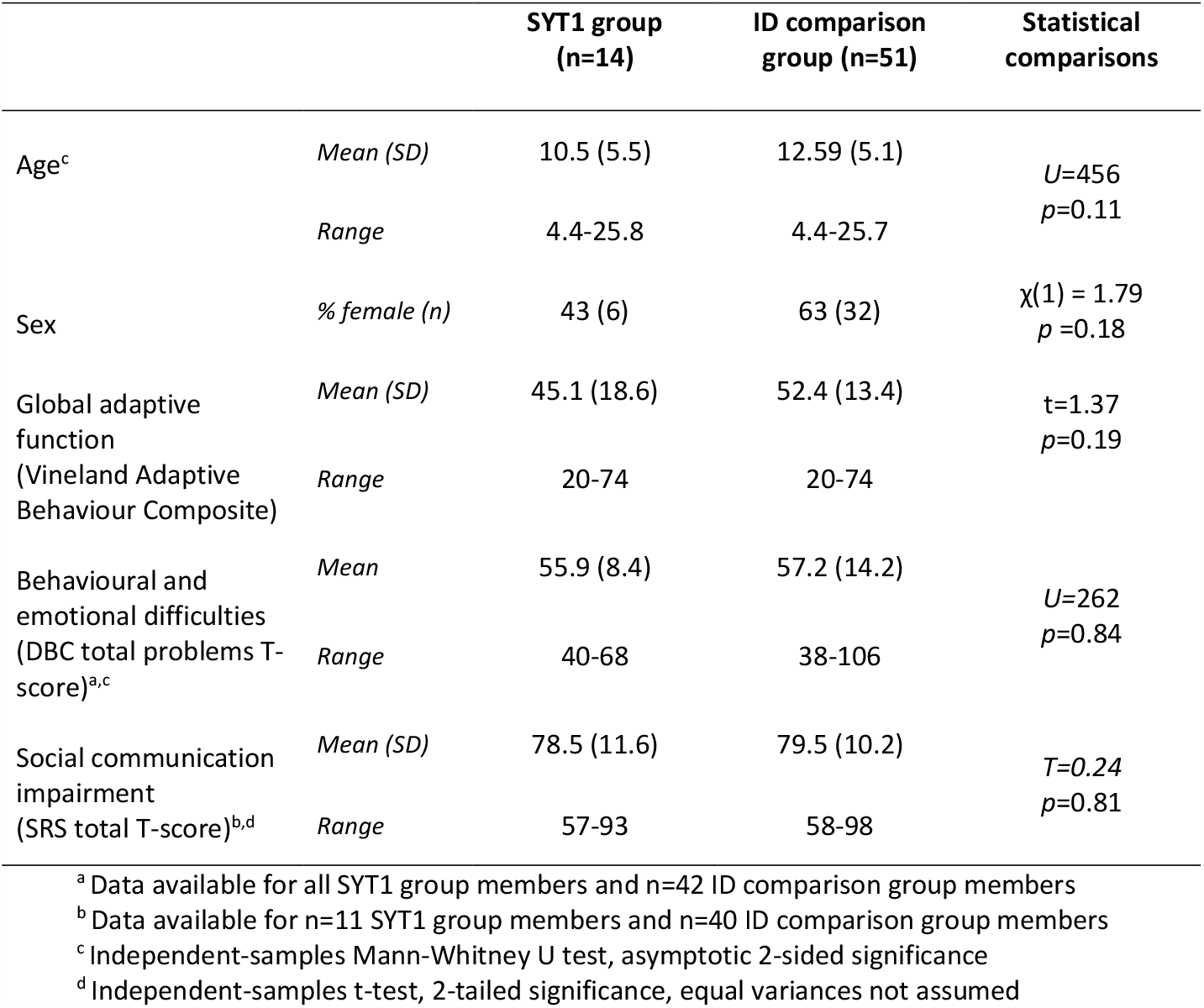
Sample characteristics for questionnaire analysis.

|  |  | SYT1 group<br>(n=14) | ID comparison<br>group (n=51) | Statistical<br>comparisons |
| --- | --- | --- | --- | --- |
| Age<sup>c</sup> | *Mean (SD)* | 10.5 (5.5) | 12.59 (5.1) | *U*=456<br>*p*=0.11 |
|  | *Range* | 4.4-25.8 | 4.4-25.7 |  |
| Sex | *% female (n)* | 43 (6) | 63 (32) | $\chi(1) = 1.79$<br>*p*=0.18 |
| Global adaptive<br>function<br>(Vineland Adaptive<br>Behaviour Composite) | *Mean (SD)* | 45.1 (18.6) | 52.4 (13.4) | *t*=1.37<br>*p*=0.19 |
|  | *Range* | 20-74 | 20-74 |  |
| Behavioural and<br>emotional difficulties<br>(DBC total problems T-score)<sup>a,c</sup> | *Mean* | 55.9 (8.4) | 57.2 (14.2) | *U*=262<br>*p*=0.84 |
|  | *Range* | 40-68 | 38-106 |  |
| Social communication<br>impairment<br>(SRS total T-score)<sup>b,d</sup> | *Mean (SD)* | 78.5 (11.6) | 79.5 (10.2) | *T*=0.24<br>*p*=0.81 |
|  | *Range* | 57-93 | 58-98 |  |

<sup>a</sup> Data available for all SYT1 group members and n=42 ID comparison group members
<sup>b</sup> Data available for n=11 SYT1 group members and n=40 ID comparison group members
<sup>c</sup> Independent-samples Mann-Whitney U test, asymptotic 2-sided significance
<sup>d</sup> Independent-samples t-test, 2-tailed significance, equal variances not assumed

**Supplementary Table 3.** Clinical descriptions of individual SYT1 cases. ***–*** *Available for download separately as Excel Spreadsheet*.

|  | L159 | T196 | E209 | E219 | M303 | D304 | S309 | N341 | Y365 | D366 | I368 | G369 | N371 |
| --- | --- | --- | --- | --- | --- | --- | --- | --- | --- | --- | --- | --- | --- |
| Variant | R | K | K | Q | K/V | G | P | S | C | E | T | D | K |
| *H. sapiens* | L | T | E | E | M | D | S | N | Y | D | I | G | N |
| *P. troglodytes* | L | T | E | E | M | D | S | N | Y | D | I | G | N |
| *M. mulatta* | L | T | E | E | M | D | S | N | Y | D | I | G | N |
| *M. musculus* | L | T | E | E | M | D | S | N | Y | D | I | G | N |
| *R. norvegicus* | L | T | E | E | M | D | S | N | Y | D | I | G | N |
| *C. familiaris* | L | T | E | E | M | D | S | N | Y | D | I | G | N |
| *F. cattus* | L | T | E | E | M | D | S | N | Y | D | I | G | N |
| *B. taurus* | L | T | E | E | M | D | S | N | Y | D | I | G | N |
| *S. scrofa* | L | T | E | E | M | D | S | N | Y | D | I | G | N |
| *G. gallus* | L | T | E | E | M | D | S | N | Y | D | I | G | N |
| *X. tropicalis* | L | T | E | E | M | D | S | N | Y | D | I | G | N |
| *D. rerio* | L | T | E | E | M | D | S | N | Y | D | I | G | N |
| *D. melanogaster* | L | T | E | D | M | D | S | N | Y | D | I | G | S |
| *C. elegans* | L | T | E | E | M | D | S | N | Y | D | L | G | N |

**Supplementary Table 4. Evolutionary conservation at sites of novel *SYT1* variants.** Sequence alignment performed with COBALT (https://www.ncbi.nlm.nih.gov/tools/cobalt/cobalt.cgi). Patient variants are shown in red, while blue highlights amino acids that are not identical to the human sequence.

**Supplementary Figure 1.**
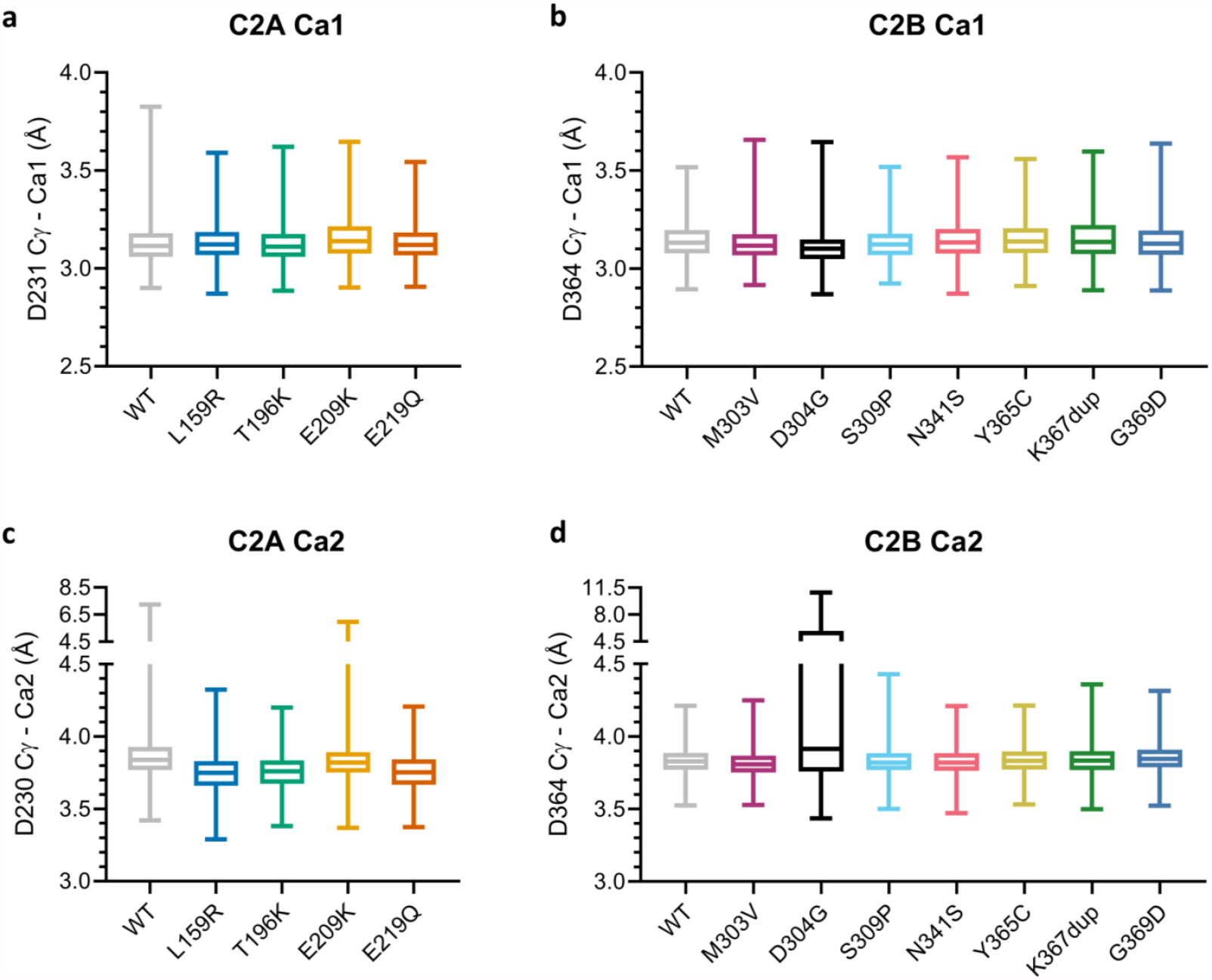
Retention of Ca^2+^ ions in binding pocket of SYT1 mutants. Ca^2+^-bound models of WT and mutant SYT1 C2A and C2B domains each underwent four ∼400ns molecular dynamics simulations. Retention of bound Ca^2+^ ions was assessed by measuring the distance between each Ca^2+^ atom and the Cγ of Arg231 of C2A (**a**,**c**) or Arg364 of C2B (**b**,**d**) in each simulation frame. Box and whisker plots show median, first and third quartiles, and minimum and maximum distances of all four trajectories per mutant. No variant substantially affected Ca^2+^ retention except for the control variant Asp304Gly, as previously shown by Baker et al. (2018)^13^.

**Supplementary Figure 2.**
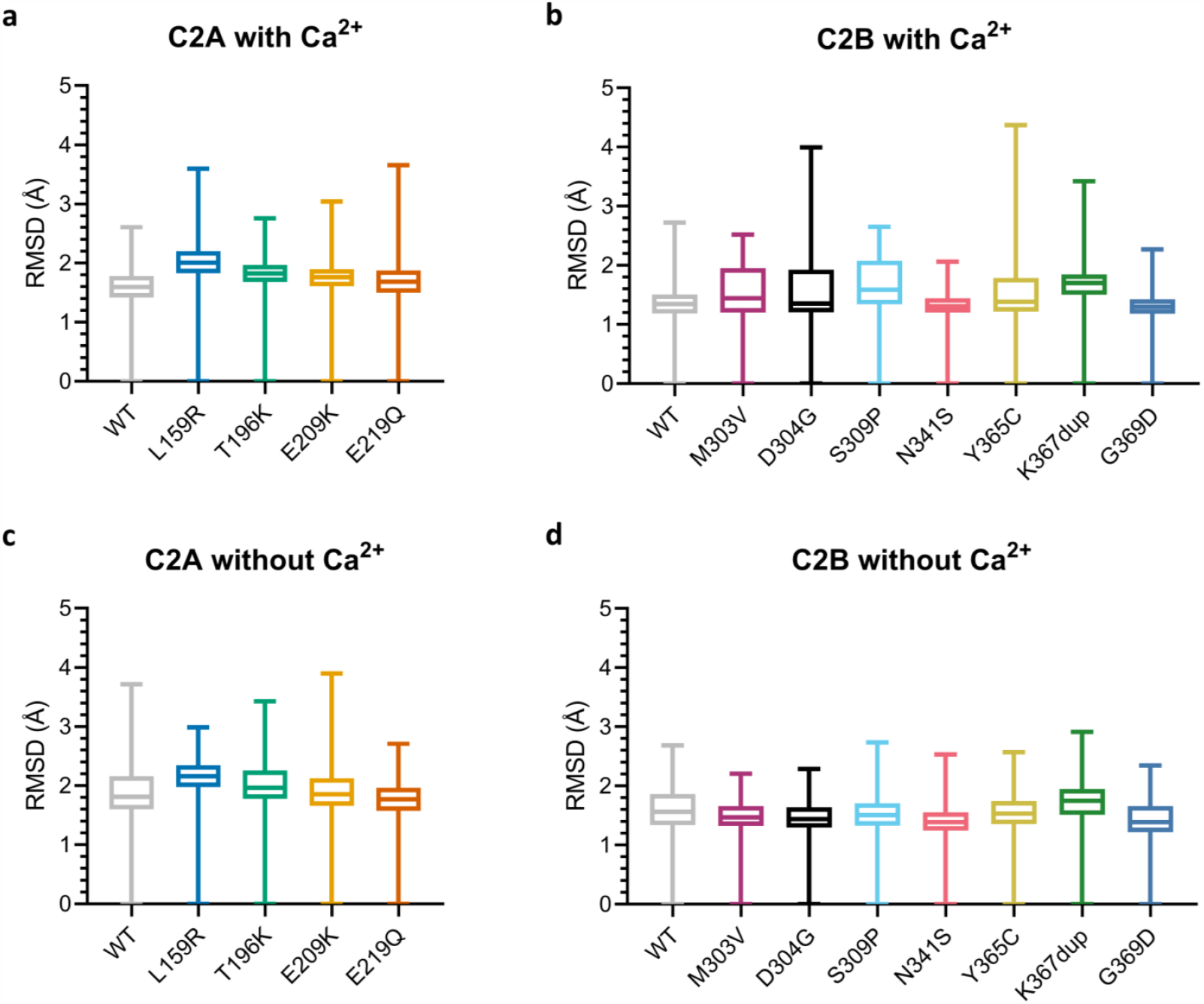
RMSD of C2A and C2B variants. Models of WT and mutant SYT1 C2A (**a**,**c**) and C2B (**b**,**d**) domains, with Ca^2+^ ions either present (**a**,**b**) or removed (**c**,**d**), each underwent four ∼400ns molecular dynamics simulations. RMSD (root-mean-square deviation) of the backbone atoms of each domain variant, compared to the starting structure, was measured over the course of each simulation. Box and whisker plots show median, first and third quartiles, and minimum and maximum RMSD values of all four trajectories per mutant.

**Supplementary Figure 3.**
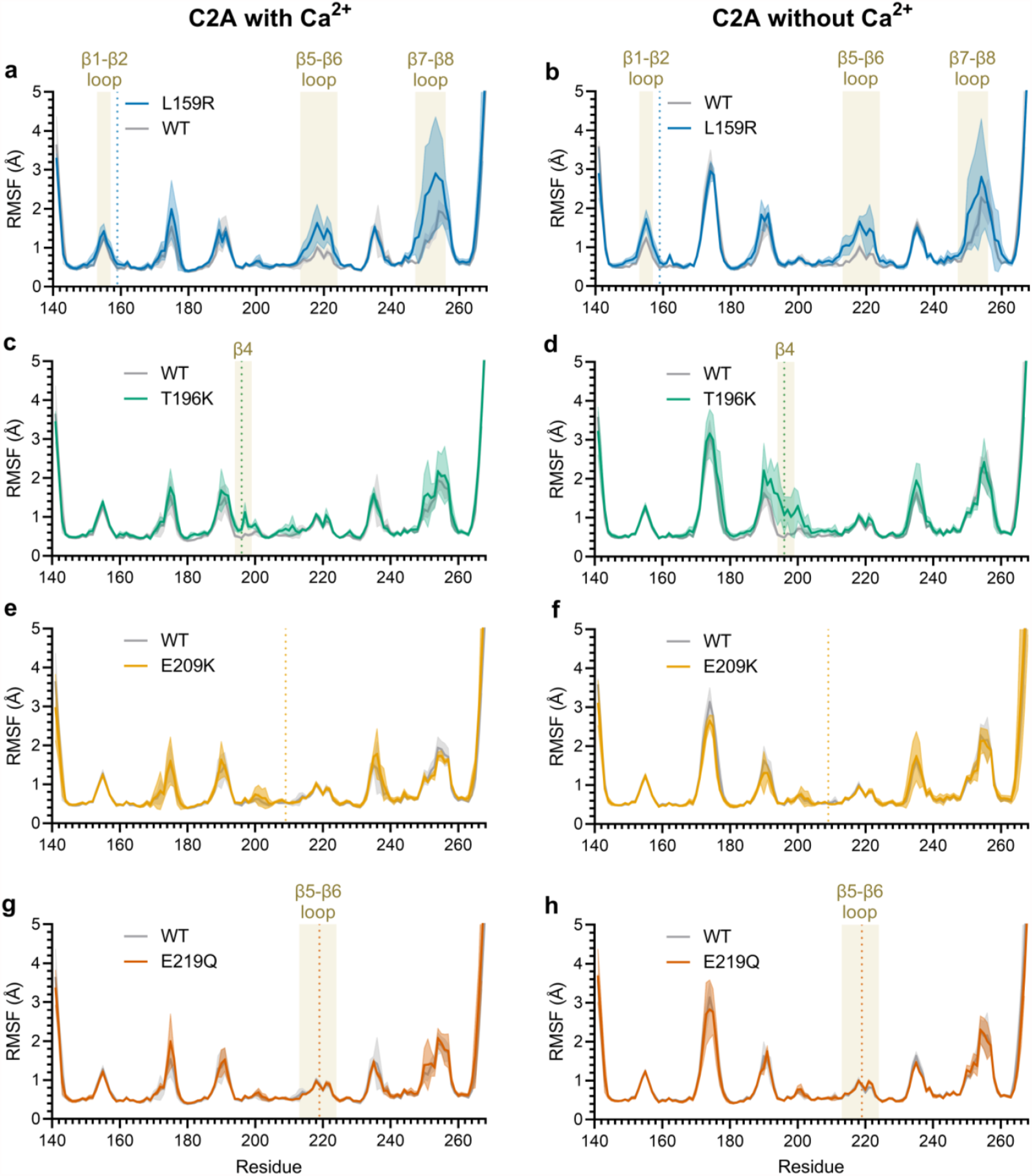
Mean RMSF traces of C2A variants. Models of WT and mutant SYT1 C2A domains, with Ca^2+^ ions either present (left) or removed (right), each underwent four ∼400ns molecular dynamics simulations. RMSF (root-mean-square fluctuations) of the backbone C-alpha atoms of each novel variant (Leu159Arg (**a**,**b**), Thr196Lys (**c**,**d**), Glu209Lys (**e**,**f**), Glu219Gln (**g**,**h**)) and WT domains were measured over the course of the simulation and plotted for each residue. Shaded regions highlight specific features of the domain as labelled (β-strands and loops between β-strands). Vertical dotted lines indicate site of mutated residue. Data are mean ± SD of four simulations.

**Supplementary Figure 4.**
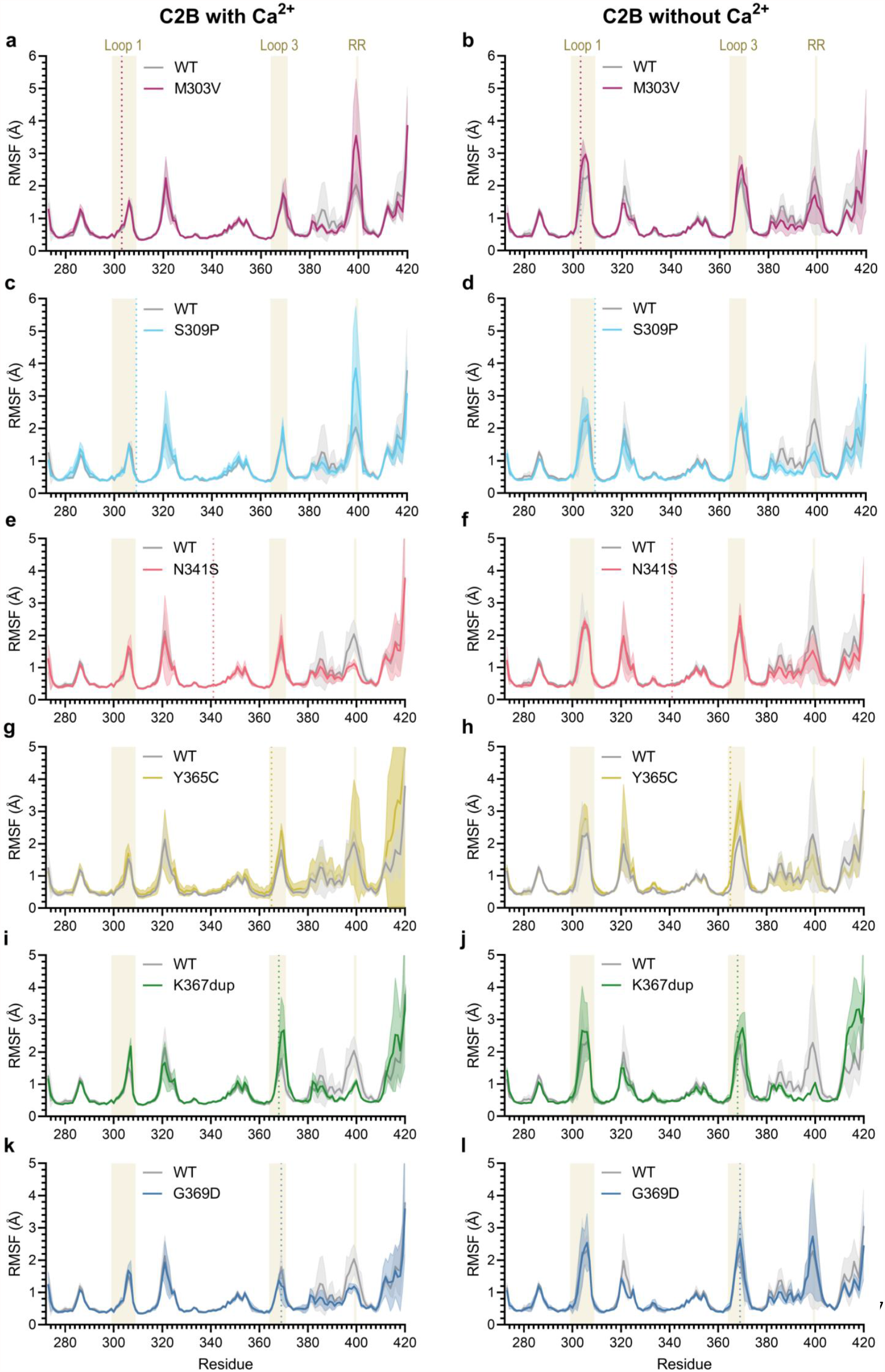
Mean RMSF traces of C2B variants. Models of WT and mutant SYT1 C2B domains, with Ca^2+^ ions either present (left) or removed (right), each underwent four ∼400ns molecular dynamics simulations. RMSF (root-mean-square fluctuations) of the backbone C-alpha atoms of each novel variant (Met303Val (**a**,**b**), Ser309Pro (**c**,**d**), Asn341Ser (**e**,**f**), Tyr365Cys (**g**,**h**), Lys367dup (**i**,**j**), Gly369Asp (**k**,**l**)) and WT domains were measured over the course of the simulation and plotted for each residue. Shaded regions highlight specific features of the domain as labelled (Ca^2+^- binding loops 1 and 3, and arginine apex (RR) i.e. Asp399, Asp400). Vertical dotted lines indicate site of mutated residue. Data are mean ± SD of four simulations.

**Supplementary Table 5.** Select intramolecular bonds altered by SYT1 mutations in molecular dynamics simulations. Values are percentage of frames in which this interaction is present, averaged across four trajectories. All amino acid numbering follows human sequence for simplicity, but note that “mutant” simulations were performed using homology models derived from rat C2A and C2B structures (rat sequence is human sequence -1). Only hydrogen bonds present in at least 10% of frames of a simulation trajectory are shown.

| Mutant | Donor | Acceptor | Bond Type | WT with Ca<sup>2+</sup> |  | Mutant with Ca<sup>2+</sup> |  | WT no Ca<sup>2+</sup> |  | Mutant no Ca<sup>2+</sup> |  |
| --- | --- | --- | --- | --- | --- | --- | --- | --- | --- | --- | --- |
|  |  |  |  | Avg | SD | Avg | SD | Avg | SD | Avg | SD |
| **E209K** | K197 backbone | E209 side chain | H-bond | 37% | 6% | - | - | 14% | 7% | - | - |
|  | K197 side chain | E209 side chain | Salt Bridge | 14% | 2% | - | - | 16% | 2% | - | - |
| **E219Q** | K223 side chain | E219 side chain | Salt Bridge | 22% | 2% | 1% | 0% | 25% | 3% | 1% | 0% |
|  | K223 side chain | E/Q219 backbone | H-bond | 13% | 4% | 10% | 2% | 12% | 2% | 11% | 2% |
|  | E/Q219 backbone | P216 backbone | H-bond | 25% | 3% | 15% | 2% | 25% | 2% | 18% | 3% |
| **S309P** | T335 backbone | S/P309 backbone | H-bond | 30% | 2% | 22% | 2% | 33% | 2% | 17% | 3% |
|  | S309 side chain | D304 side chain | H-bond | 26% | 18% | - | - | - | - | - | - |
|  | M303 backbone | S309 side chain | H-bond | 19% | 9% | - | - | 5% | 2% | - | - |
|  | S309 side chain | D364 side chain | H-bond | - | - | - | - | 12% | 15% | - | - |

**Supplementary Figure 5.**
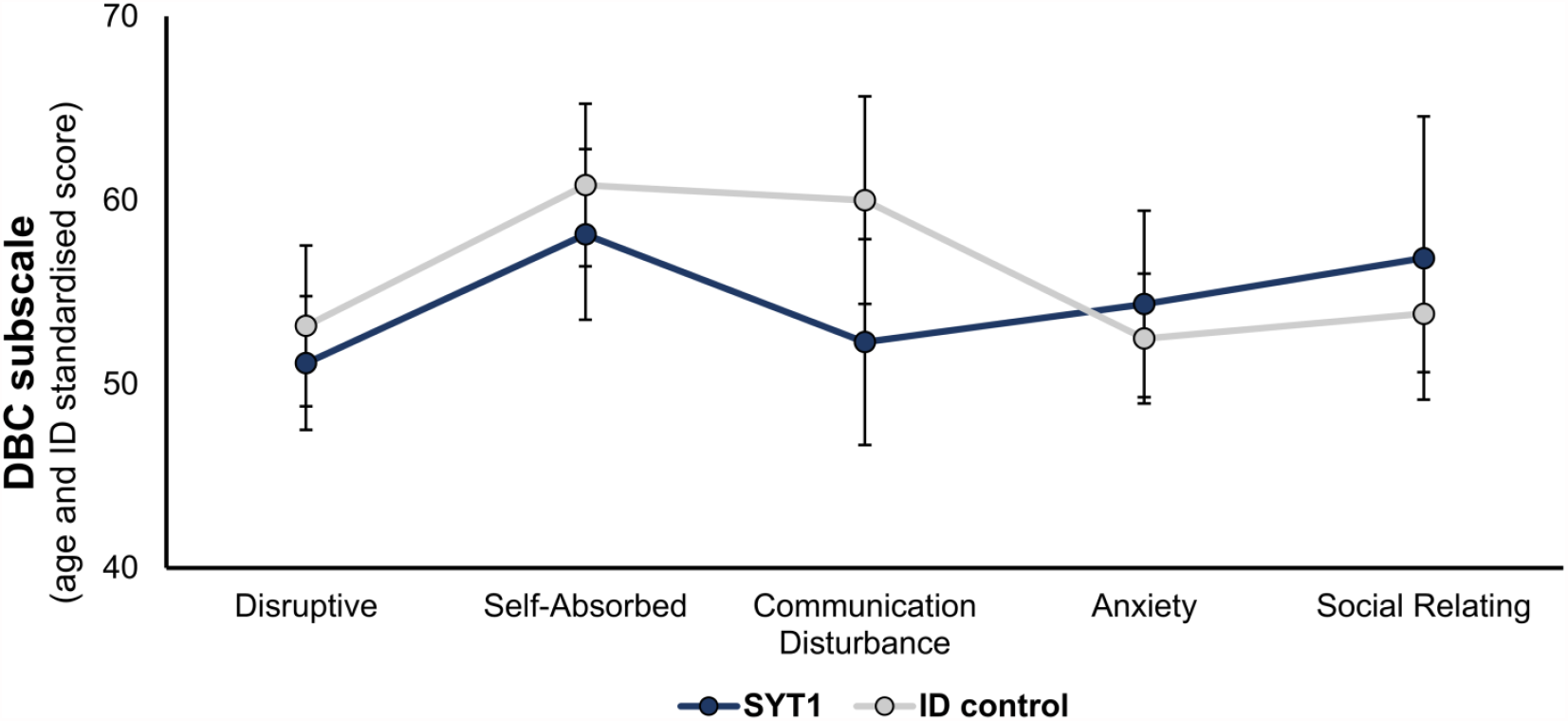
Profile of behavioural and emotional problems in SYT1 cohort. Mean scores for each subscale of the DBC (Developmental Behaviour Checklist), standardised for age and global ability, are shown for SYT1 (n=14) and ID comparison (n=42) groups. T-scores ≥ 50 indicates impairments of likely clinical concern. Error bars represent 95% confidence intervals. No significant differences between groups were observed through general linear model analysis.

**Supplementary Figure 6.**
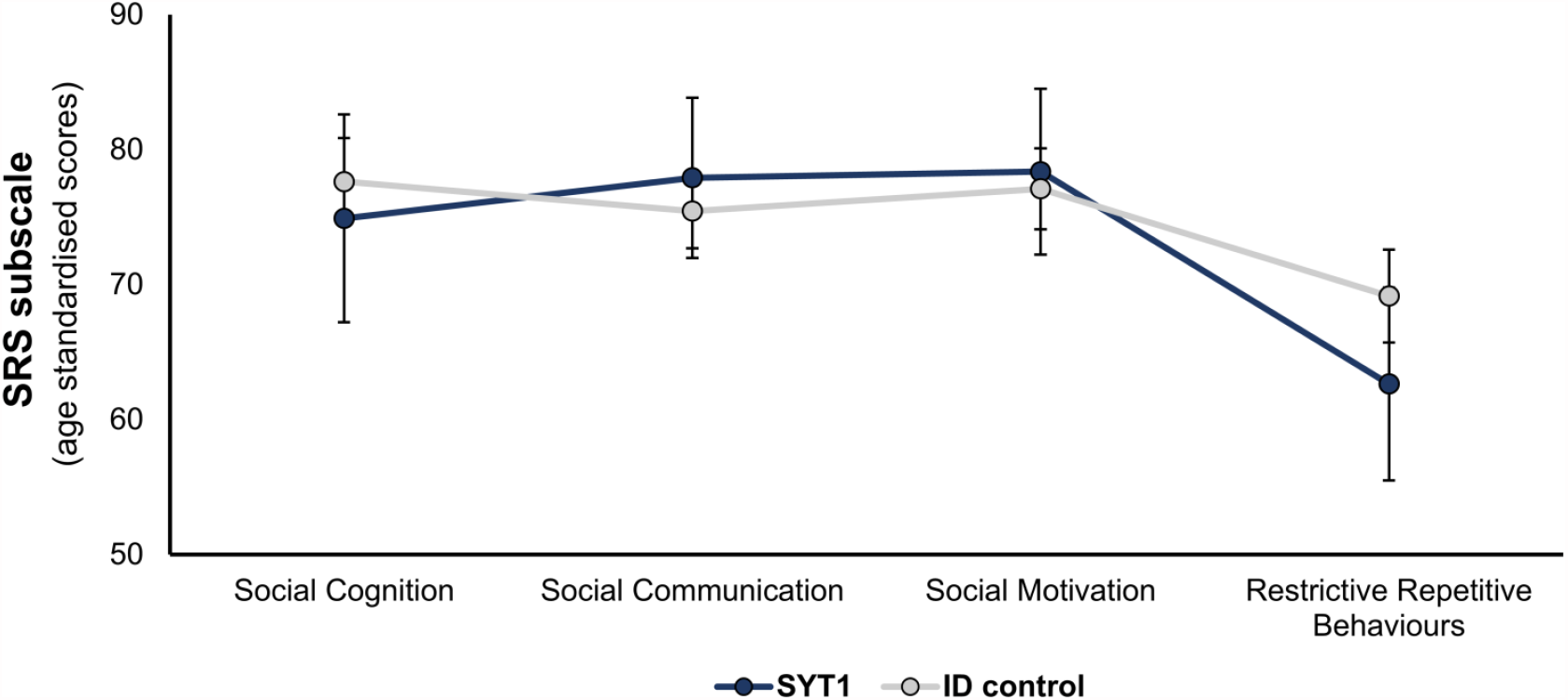
Profile of autism-related social functions in SYT1 cohort. Mean scores for each subscale of the SRS (Social Responsiveness Scale) are shown for SYT1 (n=11) and ID comparison (n=40) groups. T-score ≥ 75 indicates impairments that are suggestive of possible autism diagnosis. Error bars represent 95% confidence intervals. No significant differences between groups were observed through general linear model analysis.

**Supplementary Figure 7.**
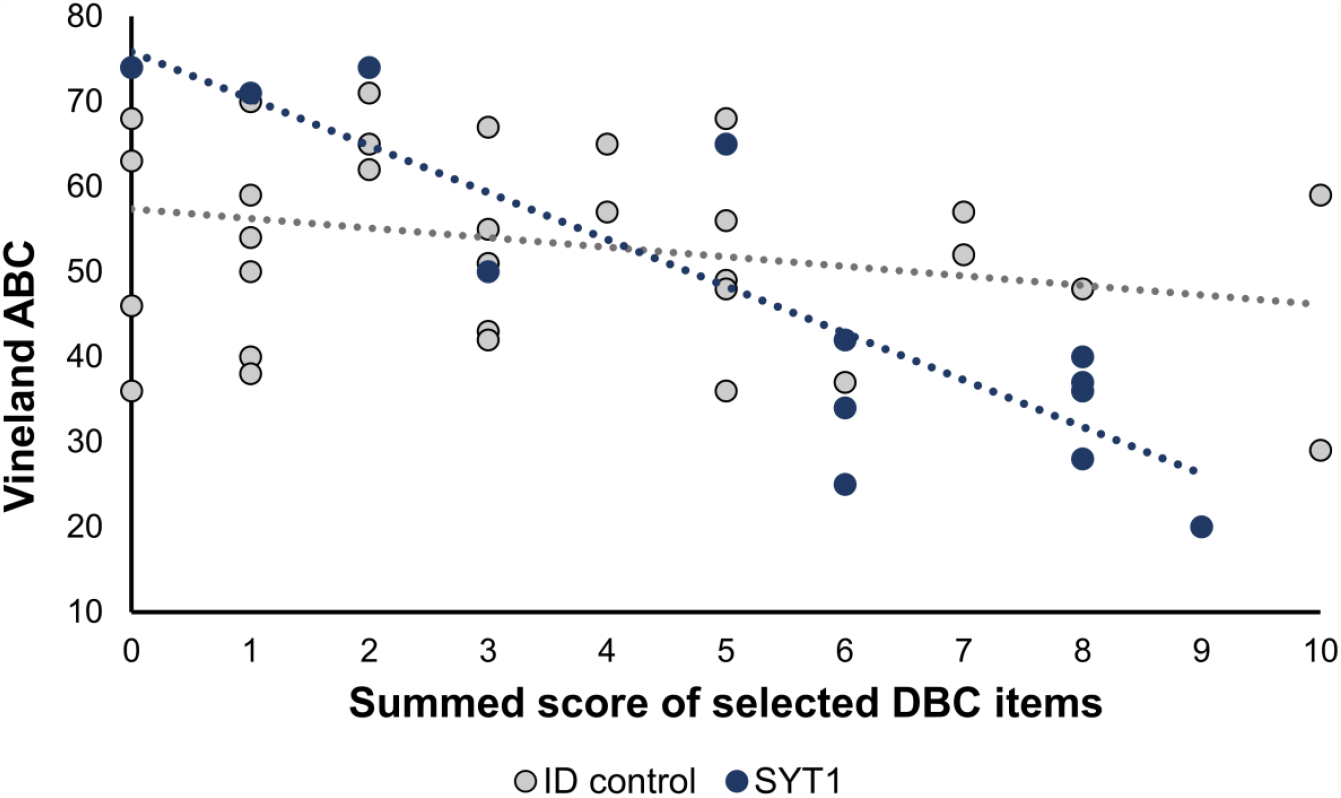
Relationship between selected DBC items and Vineland ABC in SYT1 cohort. Five items relating to self-injury, mood instability and repetitive movements were selected from the DBC-P. Raw scores (0-2) for each item were summed for each participant. Significantly higher scores on these selected items were found in the SYT1 group (n=14) compared to ID controls (n=42; p=0.02). Data show these summed scores from selected DBC items plotted against the Vineland ABC score of each individual, with each data point representing one participant. Dotted lines show linear correlation between score for selected DBC items and Vineland ABC. A significant relationship between these scores was observed only in the SYT1 group (SYT1: Spearman’s rho -0.73, p=0.003; ID controls: Spearman’s rho -0.18, p=0.26; Fisher’s test z=-2.19, p=0.01).

**Supplementary Figure 8.**
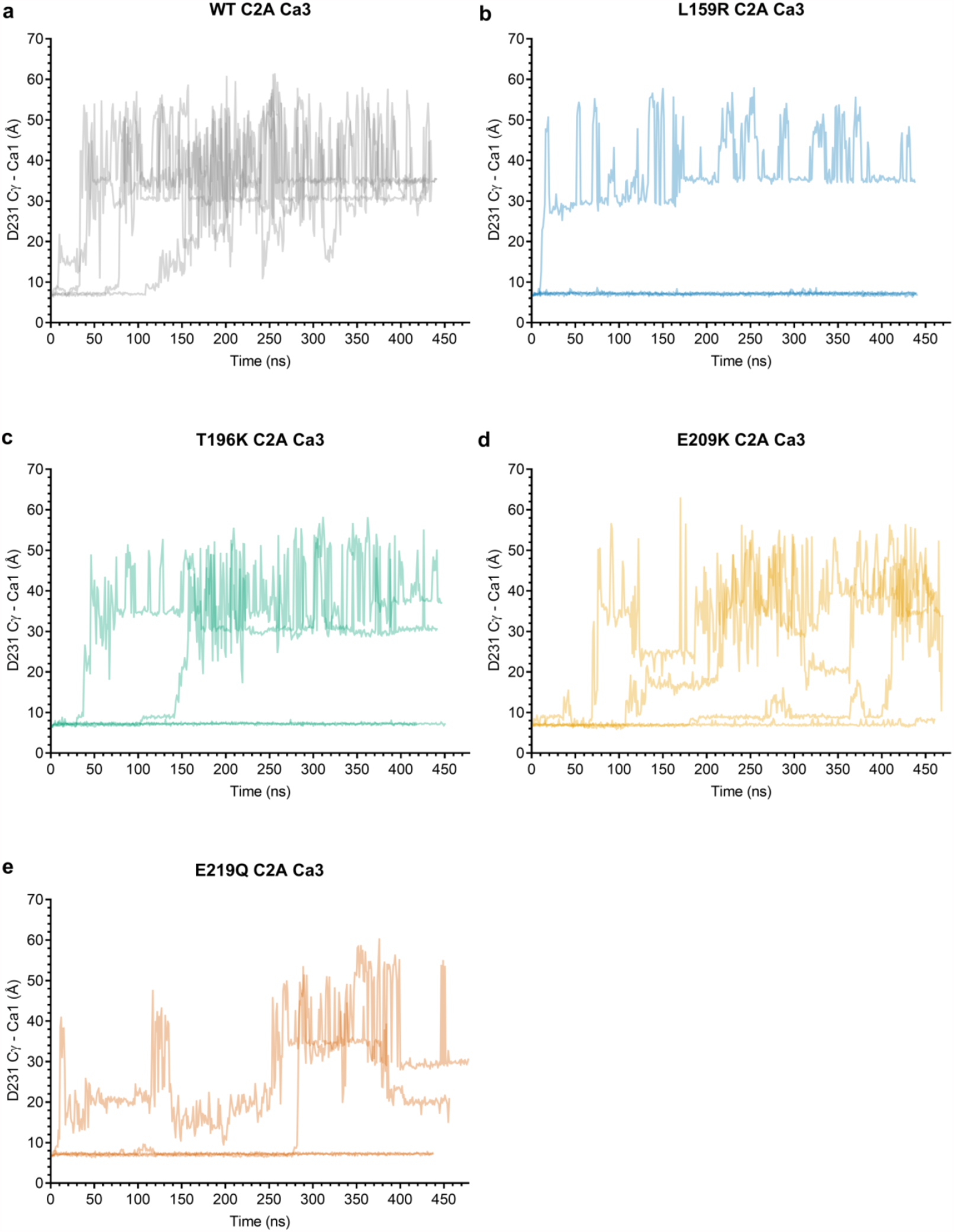
Unstable binding of Ca3 in WT and mutant C2A domains. Ca^2+^-bound models of WT and mutant SYT1 C2A domains each underwent four ∼400ns molecular dynamics simulations. Retention of bound Ca^2+^ ions was assessed by measuring the distance between each Ca^2+^ atom and the Cγ of Asp231 of C2A in each simulation frame. Data show these distances over simulation time for each of the four trajectories per mutant. (**a**) WT. (**b**) Leu159Arg. (**c**) Thr196Lys. (**d**) Glu209Lys. (**e**) Glu219Gln.

## Notes

### Competing Interest Statement

The authors have declared no competing interest.

### Author Declarations

This study was conducted within the Phenotypes in Intellectual Disability project, which received approval from Cambridge Central Research Ethics Committee (REC ref: IRAS 83633). Written informed consent was provided by each diagnosed individuals parent or consultee prior to information sharing by referring clinicians, and questionnaire completion by parents or carers.

